# An Interpretable Risk Prediction Model for Healthcare with Pattern Attention

**DOI:** 10.1101/2020.07.26.20162479

**Authors:** Sundreen Asad Kamal, Changchang Yin, Buyue Qian, Ping Zhang

## Abstract

**Background:** The availability of massive amount of data enables the possibility of clinical predictive tasks. Deep learning methods have achieved promising performance on the tasks. However, most existing methods suffer from three limitations: (i) There are lots of missing value for real value events, many methods impute the missing value and then train their models based on the imputed values, which may introduce imputation bias. The models’ performance is highly dependent on the imputation accuracy. (ii) Lots of existing studies just take Boolean value medical events (e.g. diagnosis code) as inputs, but ignore real value medical events (e.g., lab tests and vital signs), which are more important for acute disease (e.g., sepsis) and mortality prediction. (iii) Existing interpretable models can illustrate which medical events are conducive to the output results, but are not able to give contributions of patterns among medical events.

**Methods:** In this study, we propose a novel interpretable **P**attern **A**ttention model with **V**alue **E**mbedding (PAVE) to predict the risks of certain diseases. PAVE takes the embedding of various medical events, their values and the corresponding occurring time as inputs, leverage self-attention mechanism to attend to meaningful patterns among medical events for risk prediction tasks. Because only the observed values are embedded into vectors, we don’t need to impute the missing values and thus avoids the imputations bias. Moreover, the self-attention mechanism is helpful for the model interpretability, which means the proposed model can output which patterns cause high risks.

**Results:** We conduct sepsis onset prediction and mortality prediction experiments on a publicly available dataset MIMIC-III and our proprietary EHR dataset. The experimental results show that PAVE outperforms existing models. Moreover, by analyzing the self-attention weights, our model outputs meaningful medical event patterns related to mortality.

**Conclusions:** PAVE learns effective medical event representation by incorporating the values and occurring time, which can improve the risk prediction performance. Moreover, the presented self-attention mechanism can not only capture patients’ health state information, but also output the contributions of various medical event patterns, which pave the way for interpretable clinical risk predictions.

**Availability:** The code for this paper is available at: https://github.com/yinchangchang/PAVE.

## Background

With the increased growth of Electronic Health Records (EHRs) both in volume and diversity during the last decades, it becomes possible to apply clinical predictive models to improve the quality of clinical care. EHRs are temporal sequence data and consist of diagnosis codes, medications, lab results, and vital signs. Patient health information contained in the massive EHRs is extremely useful in different tasks within the medical domain, such as risk prediction [1], computational phenotyping [2], and patient similarity analysis[3]. In this paper, we focus on clinical risk prediction tasks. Most state-of-the-art clinical risk predictive models are based on deep learning, and trained in an end-to-end way. Recurrent Neural Network (RNN), a popular deep learning model for modeling sequences, has achieved good performance in clinical risk prediction tasks recently [4, 5, 6]. However, there are still some challenges in the field. (i) Most existing methods [7, 8] represent medical events as embedding vectors, which lose real value information of the medical events (e.g., lab tests and vital signs). (ii) Lab tests are diagnosis-driven and therefore EHRs have lots of missing value for lab tests. Many methods [9] impute the missing value and then train their models based on the imputed values. The models’ performance is highly dependent on the imputation accuracy. (iii) Existing interpretable models are only able to provide instance-wise variable importance (i.e., to compute each medical event’s contribution to the disease risks) rather than pattern-wise importance. It is possible that when some clinical events occur simultaneously, it may lead to a sharp increase to risk while each event alone does not cause high risk.

In this study, we propose a new interpretable **P**attern **A**ttention model with **V**alue **E**mbedding (PAVE), which is totally based on attention mechanism. For each patient, medical events, values (e.g., lab test and vital sign values) and their corresponding occurring times are represented as embedding vectors and projected to a medical semantic space. Then a self-attention layer is leveraged to capture the meaningful patterns among medical events. A pattern attention module is proposed to attend to the event patterns and produce an attention vector for each patient. Finally, we use a fully connected layer to predict a patient’s risk for future clinical outcomes. By analyzing the self-attention weights and pattern attention weights, our model is able to compute the contribution rates of various medical event patterns, thus paving the way for interpretable clinical risk predictions.

In order to demonstrate the effectiveness of the proposed PAVE, we compare our model against both traditional machine-learning methods (e.g., logistic regression, random forest) and recent deep-learning methods (e.g., RETAIN) on sepsis and mortality risk prediction tasks. We conducted experiments on both a publicly available MIMIC-III dataset [10] and our proprietary EHRs data. The experimental results show that PAVE outperforms all the baselines in both datasets and various settings, which demonstrates the effectiveness of the proposed model. Moreover, after PAVE is well trained, it is also able to find the EHRs event patterns with high contribution rates to high mortality risks. To highlight the handout of the proposed framework is as follows:

- We propose a novel interpretable risk prediction model PAVE, which is based on a self-attention mechanism and achieves better performance than the baselines.
- The presented self-attention mechanism can automatically capture meaningful patterns and is helpful to find the patterns related to high risks. To the best of our knowledge, this work is the first attempt to identify the contributions of patterns.
- We propose a new value embedding that can map values into vectors, so we don’t need to impute the missing values.
- Our medical event embedding module can take medical events’ occurring time into account.

### Related Work

Due to their promising performance in clinical risk prediction task, deep learning methods have attracted significant interest from healthcare researchers. In this section, we go through with the existing work related to deep learning models, including risk prediction, attention mechanism, and clinical models’ interpretability.

### Risk Prediction for Healthcare

Extensive research has shown the potential of early prediction of the risk of diseases from Electronic Health Records (EHRs) data, which has tempted substantial attention [11, 12, 9, 13, 14]. In this section, we mainly focus on Recurrent Neural Networks (RNN) based models. RNN can be used for patient subtyping [2], phenotyping [15], similarity measurement [3], and missing values imputation [9, 16], which are highly related to risk prediction tasks. For some RNN based approaches, the relationships between subsequent visits are usually not considered. To address the issue, Dipole [7] adopts attention mechanisms to capture the visits’ relations and therefore significantly improves the prediction accuracy.

When preprocessing the EHRs data, most existing models ignore the time intervals between neighboring medical events. However, the time intervals are common and important in many healthcare applications. Therefore, a time-aware patient subtyping model [2] is proposed to take into account time intervals in patients’ EHRs data. It is demonstrated that taking time intervals into account can significantly improve the model’s performance.

### Attention Mechanism

There are all kinds of medical events (e.g., diagnoses and medications) in EHRs data, which includes redundant and useless information. Only the events related to some specific diseases are crucial to predict risk. Therefore, attention mechanism is introduced to automatically attend to the useful events [17, 4, 7].

The attention mechanism has been shown to be helpful in the natural language processing domain. Vaswani et. al. propose Transformer [18] for machine translation task. Transformer uses self-attention to capture the relations between input words inside a sentence. The self-attention mechanism is highly parallelizable and easy to train. This work adopts a self-attention mechanism to do clinical risk prediction tasks and simultaneously aims to find clinically significant patterns related to sepsis and mortality risk with self-attention.

### Interpretability

In the clinical domain, models’ interpretability could be more important than their performance. Black-box approaches, especially deep learning methods, are not trusted by doctors and therefore not applied to real clinical situations. It motivates a lot of work focused on the interpretability of risk predictive models. RETAIN [4] is the first work that can interpret why the model makes particular predictions. It utilizes two attention modules (i.e., visit-level and code-level attention) that detect influential visits and significant medical codes. The attention weights of events indicate their importance for clinical outputs.

Then RETAIN input the weighted average of each patient’s events’ embeddings to a fully connected layer to predict the risk, which loses temporal information (e.g., the visits occurring order in patients’ EHRs data). Thus RETAIN achieves limited performance. Inspired by RETAIN, Zhang et.al. [17] propose an interpretable model to predict the risk of heart failure (IFM). IFM presents a position attention layer to capture clinical events’ order. However, IFM ignores the irregular time intervals between visits in patients’ EHRs data. Both the studies aim to calculate events’ contribution to clinical output risk, but ignore medical event patterns’ importance. It is possible that when some clinical events occur simultaneously, it may lead to a sharp increase to risk while each event alone does not cause high risk. In this study, we adopt self-attention mechanism to capture clinical significant event patterns[19].

## Methods

In this section, we give a detailed description of the proposed PAVE, which consists of four main parts. First, an embedding module represents medical events, variable values and the happening time as vectors. Then, a self-attention module is used to capture the pattern information between events. Next, a pattern attention module is followed to fuse all the pattern features, which are sent to a fully connected layer to predict the clinical outcomes. The framework of PAVE is shown in Figure 1.

**Figure 1.**
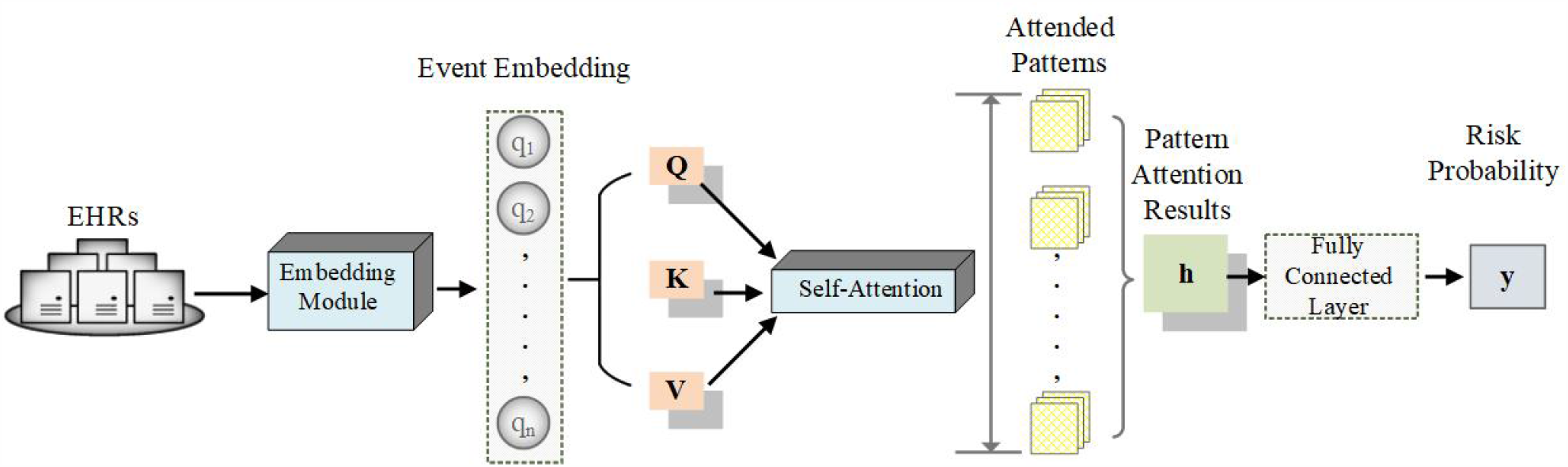
Framework of PAVE. Given a patient, the event embedding module takes his/her EHRs as inputs and generates a sequence of embedding vectors *q* =*{q*_1_, *q*_2_, …, *q*_*n*_*}*. Then three fully connected layers are followed to map *q* to queries *Q*, keys *K* and values *V*. Next, a self-attention module is adopted to attend to meaningful patterns between medical events, which are sent to a pattern attention module to generate the attention result *h*. Finally, a fully connected layer and Sigmoid layer are leveraged to output the clinical outcome risk.

### Problem Definition and Notation

The risk prediction task can be regarded as a binary classification problem. Given a sequence of medical events, the framework aims to predict if the patient will have a certain medical event (e.g., diagnosis codes, mortality) in the future.

A patient’s EHRs data consist of two main parts: static information and dynamic information. Static information is his/her demographics, such as gender and age. We represent each patient’s demographics as one-hot vectors. Patients’ ages are divided into several age groups (e.g., 20-29, 30-39).

The dynamic information is his/her historical records, including diagnosis codes, medications, lab tests, vital signs (patients in ICU have vital sign data). Each diagnosis code is Boolean-value data and others are real-value data. There could be several diagnosis codes, many collections of lab tests and vital sign data in one visit. There are usually some missing values in some items of the lab test and vital signs in each collection.

Given a patient, his/her data are denoted as (*x, ŷ*). The input data *x* includes the input demographics *d* and a sequence of *n* EHRs records, denoted as (*e*_1_, *t*_1_), (*e*_2_, *t*_2_), ‥, (*e*_*n*_, *t*_*n*_). For each event *e*_*i*_, its happening time is represented as *t*_*i*_. *ŷ* is the risk ground truth.

### Embedding Module

In this subsection, we present a new event embedding with the consideration of variable values, the corresponding happening time and patient demographics. As Shown in Figure 2, the embedding module takes event (as well as the values), happening time, demographics as input and adopts three embedding layers to project them into vectors.

**Figure 2.**
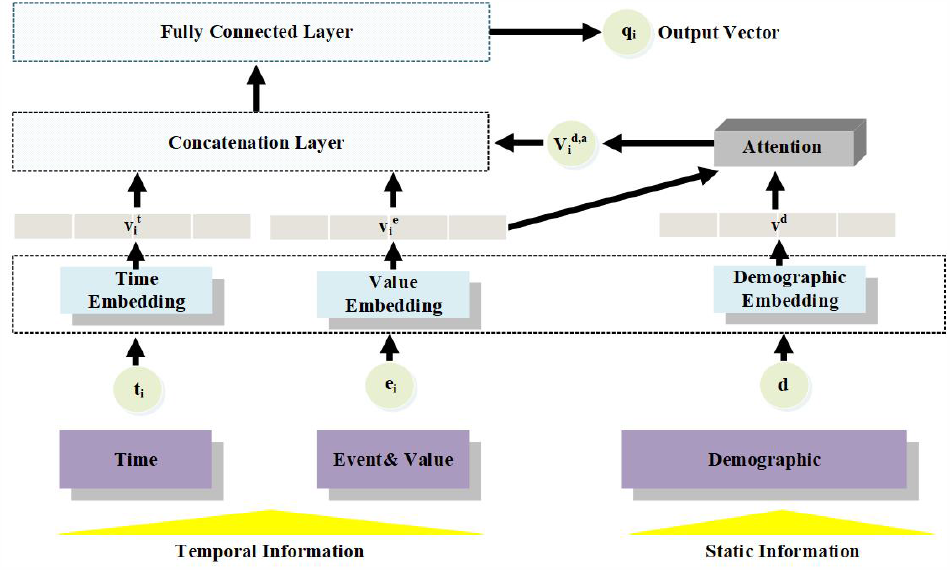
For each event *i*, we embed its happening time into vector 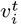, and its event value into vector 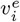. Then 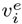 is used to attend to the patient’s demographic information. The attention result 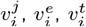 are concatenated and a fully connected layer is followed. The output vector *q*_*i*_ is the embedding vector for the event *i*.

#### Time Embedding

The first embedding layer is time embedding layer, which map the happening time *t*_*i*_ into a vector 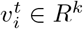. *t*_*i*_ is the interval time between the happening time of event *e*_*i*_ and the last event time. The *j*^*th*^ dimension of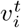 is computed as:

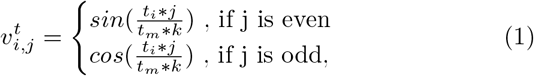

where *t*_*m*_ is the maximum of time intervals, *k* denotes the dimension of 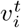.

#### Value Embedding

The second is medical event value embedding layer, which map each event *e*_*i*_ and its value *v*_*i*_ into a vector 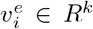. Given an event and its value, we map the event into a vector 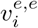 via a fully connected layer. If the event value is Boolean value (e.g., diagnosis code), we directly use 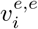 as 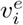. Otherwise for float value events (e.g., lab tests), given the value *v*_*i*_ of event *e*_*i*_, the value embedding layer generate a vector 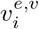 in the same way as time embedding layer. The *j*^*th*^ dimension of 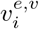 is computed as:

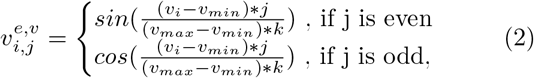

where *v*_*min*_ and *v*_*max*_ are the minimum and maximum values of the corresponding variable, *k* denotes the dimension of 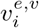. Given 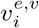and 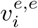, a linear function is used to combine them to 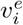:

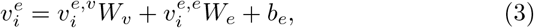

where *W*_*v*_, *W*_*e*_ ∈ *R*^*k×k*^ and *b*_*e*_ ∈ *R*^*k*^ are learnable parameters.

#### Demographic Embedding

The third embedding layer is demographic embedding layer, which embeds *d* into a matrix *v*^*d*^ *∈R*^|*d*|*×k*^. A demographic attention mechanism is leveraged to attend to the demographic information.

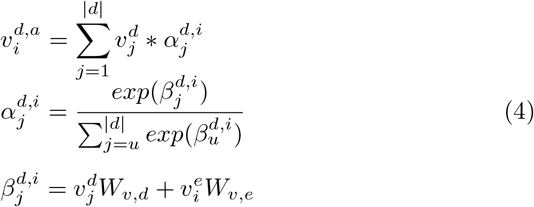

, where *W*_*v,d*_, *W*_*v,e*_ *∈ R*^*k*^ are learnable parameter, 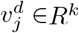 denotes the *j*^*th*^ dimension of 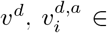 is the demographic attention result.

Given the embedding and attention results (i.e., 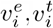 and 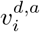), using a concatenation operation and a fully connected layer, the *i*^*th*^ event and the patient’s demographics are projected into an embedding vector *q*_*i*_ *∈ R*^*k*^.

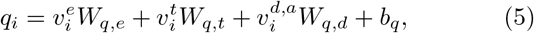

where *W*_*q,e*_, *W*_*q,t*_, *W*_*q,d*_ *∈ R*^*k×k*^ and *b*_*q*_ ∈ *R*^*k*^ are learnable parameters.

### Self Attention Module

Given a patients, his/her sequence of final embeddings of events *q* = *{q*_1_, *q*_2_, …, *q*_*n*_*}* are input to self-attention module to capture useful patterns between related events. Three fully connected layers are used to map *q* into three matrices *Q, K, V ∈ R*^*n×k*^. Then, we compute the dot products of each query *Q*_*i*_ with other keys *K*_*j*_ and calculate the attention weight *α*_*ij*_ with a softmax function. Obtaining the weight, the sum of query event’s value and attention result of key events’ values is output as the pattern attention out-come *P ∈R*^*n×k*^. The *i*^*th*^ dimension *P* is computed as follows:

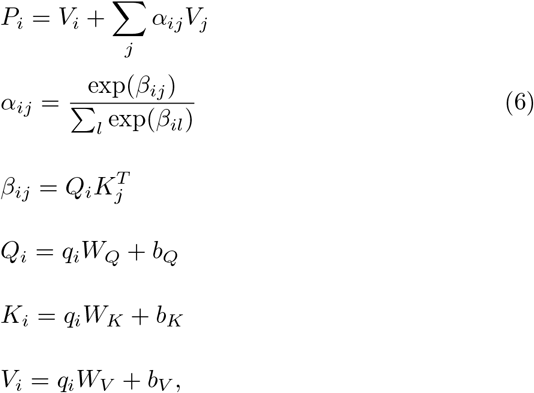

where *W*_*Q*_, *W*_*K*_, *W*_*V*_ ∈ *R*^*k×k*^ and *b*_*Q*_, *b*_*K*_, *b*_*V*_ ∈ *R*^*k*^ are learnable parameters. Given two events *i* and *j*, the product between query *Q*_*i*_ and key *K*_*j*_ represents their relevance *beta*_*ij*_. A softmax layer is followed to generate the attention weight *α*_*ij*_. Finally, a soft attention layer is used to produce the pattern vector *P*_*j*_. The self-attention module can capture two-event patterns. By stacking more self-attention layer, PAVE also has the potential to capture more complex medical patterns with more events.

### Pattern attention Module

There are various patterns in each patient’s EHRs data, only some are useful for risk prediction goal. Given the pattern embeddings *P ∈ R*^*n×k*^, a pattern attention mechanism is used to attend to the meaningful patterns.

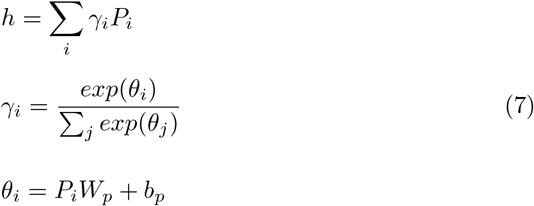

, where *W*_*p*_ *∈ R*^*k*^ and *b*_*p*_ *∈ R* is learnable parameters. Given a medical event pattern *i* for a patient, a fully connected layer is adopted to compute its relevance *θ*_*i*_ to the risk prediction task. Then a softmax layer is followed to compute the weights for different patterns. Finally a soft attention is used to combine various patterns and produce a vector *h*, which contains the patient’s clinical risk information.

### objective function

A fully connected layer and sigmoid layer are followed to predict the risk probability:

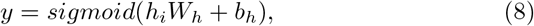

where *W*_*h*_ *∈ R*^*k*^ and *b*_*h*_ *∈ R* are learnable parameters. The cross-entropy between ground truth *ŷ* and predicted result *y* is used to compute loss:

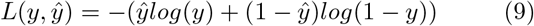

### Interpretability

The interpretability is that PAVE can compute each pattern’s contribution to the output. Given pattern (*i, j*), including event *i* and event *j*, the contribution *C*_*ij*_ is calculated as follows:

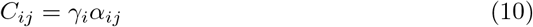

## Results and Discussion

In order to evaluate the effectiveness of the proposed PAVE, we compare our model with some state-of-art methods on two real-world clinical datasets: publicly available MIMIC-III [10] and a proprietary EHRs database. The experiments are conducted on two different tasks: sepsis onset prediction and mortality prediction.

### Datasets

Both the datasets of sepsis prediction and mortality prediction tasks are from Intensive Care Unit (ICU).

#### Sepsis prediction

The first dataset is extracted from a real-world proprietary EHRs database. We use patients’ demographics information and 28 kinds of time series features including vital signs and lab tests to predict sepsis onset after several hours. Sepsis is one of the leading causes of mortality in hospitalized patients. We follow the sepsis 2 definition [20, 21]. The sepsis 2 patients must meet at least two of the following four SIRS criteria:

- Body temperature *>* 38.0 or *<* 35.0
- Respiratory rate *>* 20 or PaCO2 *<* 32mmHg
- Heart rate *>* 90/min
- WBC *>* 12k or *<* 4k or Band *>* 10%

#### Mortality prediction

The second dataset is publicly available dataset MIMIC-III [10]. We use patients’ demographics and 8 vital signs data to predict the mortality in the coming hours. For each case patient (with sepsis 2 onset or mortality) on both datasets, 3 patients with the same age and gender are chosen as the controls. For both cases and controls, our model predicts whether the patients suffer from sepsis onset or mortality after a hold-off prediction window (e.g., 10, 8, 6, 4 hours). PAVE and baselines take patients’ observed variables during the last 48 hours as inputs (the data in the hold-off windows are excluded). The statistics of the selected datasets are listed in Table 1. The selected variables are listed in Table 2.

**Table 1.** Statistics of Datasets. Note that AVG No. of events and AVG No. of collections are calculated based on the last 48 hours data for each patient.

|  | Sepsis | Mortality |
| --- | --- | --- |
| No. of case patients | 10000 | 5000 |
| No. of control patients | 30000 | 15000 |
| No. of Male/Female | 21628/18372 | 10720/9280 |
| AVG age | 62.7 | 68.4 |
| No. of unique events | 28 | 8 |
| AVG No. of events | 225.8 | 190.9 |
| AVG No. of collections | 43.7 | 34.1 |

**Table 2.** Selected Variables use for sepsis onset and mortality prediction.

| Variables | Sepsis | Mortality |
| --- | --- | --- |
| Anion Gap | ✓ |  |
| Blood Urea Nitrogen | ✓ |  |
| Braden Scale | ✓ |  |
| Chlorine | ✓ |  |
| Creatinine | ✓ |  |
| CO2 | ✓ |  |
| Diastolic Blood Pressure | ✓ | ✓ |
| FiO2 | ✓ |  |
| Glasgow Coma Score | ✓ |  |
| Glucose | ✓ | ✓ |
| Heart Rate | ✓ | ✓ |
| Hct | ✓ |  |
| Hgb | ✓ |  |
| MAP | ✓ |  |
| MCH Concentration | ✓ |  |
| Mean Blood Pressure | ✓ | ✓ |
| Pain Score | ✓ |  |
| Platelet | ✓ |  |
| Potassium | ✓ |  |
| Pulse | ✓ |  |
| RBC | ✓ |  |
| Respiratory Rate | ✓ | ✓ |
| SO2 | ✓ |  |
| sodium | ✓ |  |
| SPO2 | ✓ | ✓ |
| Systolic Blood Pressure | ✓ | ✓ |
| Temperature | ✓ | ✓ |
| WBC | ✓ |  |

### Methods for Comparison

To validate the performance of PAVE, we compare it with the following models, including three traditional machine learning methods and four deep learning methods. In order to demonstrate the effectiveness of the proposed time embedding and event embedding, we also implement three versions of PAVE.

### Random Forest (RF)

We represent each patient’s demographics into a vector. For each variable, we extract the minimum and maximum value. The concatenation vectors of the values of patients are used to train the Random Forest model.

### Logistic Regression (LR)

We train the logistic regression model with the same vectors as random forest. The logistic regression is trained with five various solvers, including *lbfgs, new-cg, liblinear, sag* and *saga*. We choose the solver with the best performance in validation set.

### Support Vector Machine (SVM)

We train the support vector machine model with the same vectors as random forest. The support vector machine is trained with four different kernels, including *poly, rbf, linear* and *sigmoid*. The kernel with the best performance in the validation set is used to predict the risk in the test set.

### GRU and LSTM

GRU [22] and LSTM [23] are classical RNN based models, which both introduce various gates to improve RNN’s performance.

### RETAIN

The REverse Time AttentIoN model (RETAIN) [4] is the first work that tries to interpret model’s disease risk prediction results with two attention modules. The attention modules generate weights for every medical event. The weights are helpful to analyze different events’ contributions to the output risk.

### IFM

IFM [17] is an interpretable heart failure risk prediction model, which is also based on attention mechanism and leverages the attention weights to interpret the outputs. In this work, we modify the IFM to predict sepsis onset and mortality. **PAVE**^*−T*^ : PAVE^*−T*^ removes the time embedding module when predicting patient risks.

### PAVE^−V^ : PAVE^−V^

removes the variable value embedding. The method prefills the missing values with mean values and takes the prefilled values as inputs but not the value embeddings.

### PAVE

PAVE is the main version of the proposed model.

### Implementation Details

We implement all the baselines and our proposed PAVE models with PyTorch 0.4.1^[1]^. For training models, we use Adam optimizer with a mini-batch of 64 patients. We train on 1 GPU (TITAN XP) for 50 epochs, with a learning rate of 0.0001. We randomly divide the datasets into 10 sets. All the experiment results are averaged from 10-fold cross-validation, in which 7 sets are used for training every time, 1 set for validation and 2 sets for test. The validation sets are used to determine the best values of parameters in the training iterations. We use the area under the receiver operating characteristic curve (AUROC) in the test sets as a measure for comparing the performance of all the methods in two datasets. We use 512-dimensional embeddings to represent entities. The dimensions of query (*Q*), key (*K*) and value (*V*) matrices used in the self-attention layer are set as 128. We only use 1 layer of self-attention operation to capture two-event patterns. The number of our model’s parameters is about 5M. We use BCELoss as loss function.

## Results of Risk Prediction

As is shown in Table 3, the proposed model PAVE outperforms all the baselines, which demonstrates the effectiveness of our model.

**Table 3.** AUROC on sepsis and mortality prediction

|  | Sepsis Prediction |  |  |  | Mortality Prediction |  |  |  |
| --- | --- | --- | --- | --- | --- | --- | --- | --- |
|  | 10h | 8h | 6h | 4h | 10h | 8h | 6h | 4h |
| LR | 0.7125 | 0.7387 | 0.7439 | 0.7526 | 0.8027 | 0.8203 | 0.8428 | 0.8659 |
| RF | 0.7258 | 0.7406 | 0.7592 | 0.7618 | 0.8129 | 0.8293 | 0.8559 | 0.8731 |
| SVM | 0.7014 | 0.7539 | 0.7608 | 0.7628 | 0.8018 | 0.8352 | 0.8533 | 0.8627 |
| GRU | 0.7475 | 0.7612 | 0.7659 | 0.7673 | 0.8364 | 0.8420 | 0.8609 | 0.8823 |
| LSTM | 0.7496 | 0.7633 | 0.7674 | 0.7698 | 0.8566 | 0.8618 | 0.8734 | 0.8843 |
| RETAIN | 0.7414 | 0.7562 | 0.7578 | 0.7594 | 0.8476 | 0.8604 | 0.8747 | 0.8949 |
| IFM | 0.7458 | 0.7601 | 0.7627 | 0.7650 | 0.8657 | 0.8734 | 0.8817 | 0.8922 |
| PAVE<sup>-V</sup> | 0.7536 | 0.7682 | 0.7688 | 0.7704 | 0.8864 | 0.8863 | 0.8973 | 0.9153 |
| PAVE<sup>-T</sup> | 0.7559 | 0.7697 | 0.7703 | 0.7736 | 0.8892 | 0.8919 | 0.9032 | 0.9185 |
| PAVE | **0.7632** | **0.7727** | **0.7782** | **0.7805** | **0.8970** | **0.9041** | **0.9126** | **0.9222** |

The deep learning approaches outperform the traditional machine-learning approaches that take vectors as inputs but not sequence data. Traditional machine-learning approaches’ inputs lose the temporal information of EHR data, which are very important in the risk prediction tasks, while deep learning models are good at modeling temporal data. Thus, the deep learning baselines achieves better performance. Among the deep learning baselines, attention-based models (i.e., RETAIN and IFM) perform better than other models in the mortality prediction task, while LSTM and GRU perform better in the sepsis onset prediction task. We speculate that mortality is easier to predict based on several vital sign features, such as heart rate and respiratory rate in recent hours. Attention-based models do well in capturing important events and thus achieves better performance. Sepsis is a complex disease that is more difficult to be predicted than mortality. The prediction of sepsis onset is related to changes in patients’ health states during a relatively longer period.

LSTM and GRU are better at modeling the long time changes of the states, while RETAIN and IFM lose some temporal information with the attention mechanisms. In the clinical domain, models’ interpretability could be more important than their performance. Thus, the interpretable risk prediction models (i.e., PAVE, RETAIN and IFM) are more suitable for real-world clinical applications. Compared with RETAIN and IFM, PAVE leverages attention mechanism to focus on important events, and incorporates time information with time embedding, so it outperform RETAIN and IFM by 1.5 percent and 3 percent for sepsis and mortality prediction tasks respectively.

Among the three versions of the proposed model, PAVE^*−T*^ performs worse than PAVE, which means that with the time embedding, PAVE can capture more time information of time intervals. PAVE also outperforms PAVE^*−E*^, which takes the imputed values as inputs, but not value embeddings. The imputation strategy may introduce bias and thus be harmful to the final risk prediction tasks.

### Medical Event Pattern Analysis

PAVE is able to analyze the patterns’ contributions to the prediction. We compute each pattern’s contribution to the risk of mortality for each patient according to Eq. (10). For each variable, their values are divided into five ranges. By comparing each item value to its normal range, the item value is mapped into three ranges (e.g., low, normal and high). Then the high-value range is divided into two parts (i.e., high and very high) by comparing the value to the median of all the high values. The low-value range is divided in the same way. We display the top 10 patterns with the highest average contribution rates among all the case patients to mortality (10-h mortality prediction) in Table 4. The patterns are verified by clinicians to be high-risk signals to mortality, which demonstrate PAVE can find useful patterns in the prediction tasks.

We conducted the experiments lots of times and found some patterns always have relatively high weights. For example, the weight of the pattern (very high temperature and very low respiratory rate) is always much higher than other random patterns, which is consistent with clinical knowledge that the patients with very high temperature and very low respiratory rate simultaneously have high risk of mortality.

**Table 4.** Top 10 patterns with the highest average contribution rates (**AVG-CR**) to mortality. Note SysBP, DiasBP and MeanBP denote systolic blood pressure, diastolic blood pressure and mean blood pressure respectively.

| EVENT 1 | EVENT 2 | AVG-CR |
| --- | --- | --- |
| very low respiratory rate | very high temperature | 10.1% |
| very low respiratory rate | high SysBP | 9.5% |
| high temperature | high SysBP | 8.7% |
| very low temperature | very low MeanBP | 8.3% |
| low temperature | high MeanBP | 6.8% |
| very high SysBP | low spo2 | 6.5% |
| very high SysBP | very low DiasBP | 5.9% |
| very high respiratory rate | very high heart rate | 5.3% |
| high DiasBP | very high MeanBP | 5.2% |
| very high SysBP | high heart rate | 5.0% |

### Case Study

We applied PAVE to predict the mortality risk of a patient from the test set, who suffered mortality after 10 hours. We display the observed variables during the last 24 hours in observation window in Figure 3. RETAIN is also used to predict the mortality risk for comparison. Both PAVE and RETAIN accurately predict the patient’s mortality after 10 hours. In this case study, we mainly focus on the interpretability of the detected medical events or patterns with high contribution risks. The black stars in Figure 3 represent observed abnormal values with high instance-wise contribution risks generated by RETAIN, while the colored squares are medical event patterns detected by PAVE. In the case, PAVE found three patterns with high contribution risks: (i) high SysBP and high temperature in orange squares; (ii) high heart rate and high temperature in red squares; (iii) stable high heart rate and high respiratory rate in blue squares. The events sharing the same colors are detected patterns. Note that only the patterns with relatively high contribution risks are shown in the figure. The sizes of black stars and colored squares denote the corresponding values of contribution risks. Both the models successfully detect some crucial medical events related to high mortality risks, such as high heart rates and high temperature. PAVE focuses much more on the observed variables during the last 10 hours in the observation window (e.g., the stable high heart rate and high respiratory rate in blue squares), while RETAIN attends to lots of earlier events but ignore the latter high heart rate and high respiratory rate in the last three collections. It means PAVE learn an knowledge that both the latter medical events and the abnormal values are more useful for accurate mortality prediction, while RETAIN only focuses on abnormal values. Moreover, when some crucial patterns (e.g., high heart rate and high respiratory rate in blue squares) appear, PAVE assigned more attention weights to the patterns than RETAIN (the colored squares have bigger size than the corresponding stars), which demonstrate that PAVE are effective for mining relative and important patterns, and pay more attention to the meaningful patterns.

**Figure 3.**
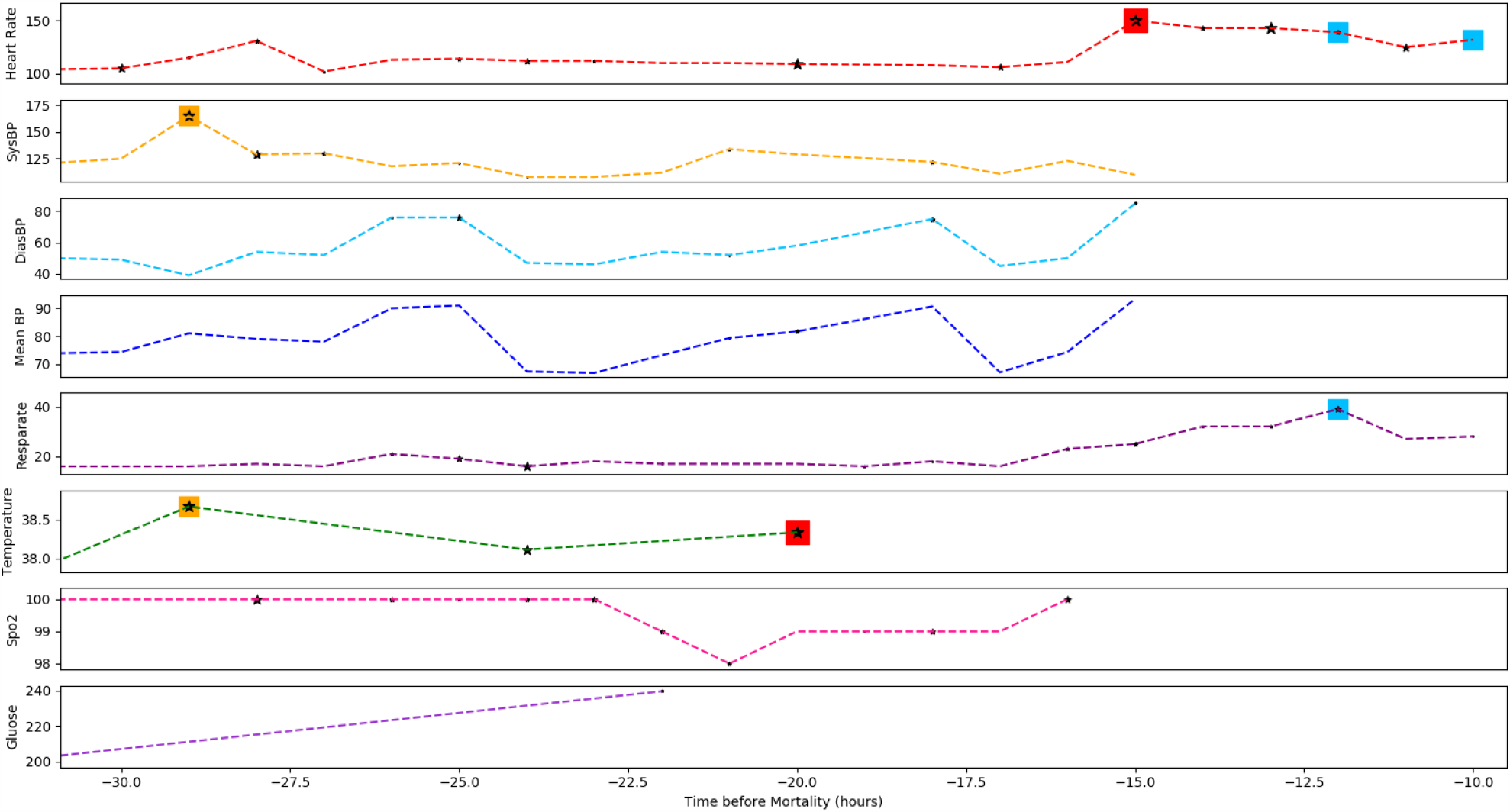
Case Study. The figure shows the observed variables of a case patient during the last 24 hours observation window (the hold-off window is 10 hours). The black stars represent observed abnormal values with instance-wise contribution risks generated by RETAIN, while the colored squares are medical event patterns detected by PAVE. The events sharing the same colors are detected patterns. The sizes of black stars and colored squares denote the corresponding values of contribution risks. Note that only the events or patterns with relatively higher contribution risks are marked in the figure. PAVE found three high-risk patterns for the patient: (i) high SysBP and high temperature in orange squares; (ii) high heart rate and high temperature in red squares; (iii) stable high heart rate and high respiratory rate in blue squares.

## Conclusion

In this work, we proposed PAVE, an interpretable pattern attention model with value embedding to predict disease risk. PAVE takes into account real-value medical events (e.g., lab tests and vital signs) by embedding the values into vectors, and therefore does not need to impute the missing values. Moreover, PAVE is based on attention mechanisms and the attention weights can be used to interpret the model’s clinical outputs. To the best of our knowledge, PAVE is the first interpretable deep learning model that can provide medical pattern-wise interpretability but not only instance-wise interpretability. Event patterns may cause a much higher risk than each single event in the pattern. We conducted expensive experiments on two real-world datasets and PAVE achieved better performance than state-of-art models. Moreover, the experimental results show that PAVE is able to detect lots of medical event patterns with high contribution rates to mortality and sepsis onset, which paves the way for interpretable clinical risk predictions.

## Data Availability

We used ublicly available dataset MIMIC-III.

https://mimic.physionet.org/

## Acknowledgements

Not applicable.

## Author’s contributions

BQ and PZ conceived the project. SK and CY developed the method. SK and CY conducted the experiments. SK, CY, BQ, and PZ wrote the manuscript. All authors read and approved the final manuscript.

## Funding

Not applicable.

## Availability of data and materials

MIMIC-III database analyzed in the study is available on PhysioNet repository. The source code is provided for reproducing and is available at https://github.com/yinchangchang/PAVE.

## Ethics approval and consent to participate

Not applicable.

## Consent for publication

Not applicable.

## Competing interests

PZ is the member of the editorial board of BMC Medical Informatics and Decision Making. The authors declare that they have no other competing interests.

## Footnotes

[1] https://pytorch.org/

## Notes

### Competing Interest Statement

The authors have declared no competing interest.

### Author Declarations

This study uses the MIMIC-III dataset. We are using the MIMIC IRB. This study was approved by the Institutional Review Boards of Beth Israel Deaconess Medical Center (Boston, MA, USA), the Massachusetts Institute of Technology (Cambridge, MA, USA) and Institute for Infocomm Research (Singapore). Requirement for individual patient consent was waived because the study did not impact clinical care and all protected health information was de-identified. De-identification was performed in compliance with Health Insurance Portability and Accountability Act (HIPAA) standards in order to facilitate public access to MIMIC-III. Deletion of protected health information (PHI) from structured data sources (e.g., database fields that provide patient name or date of birth) was straightforward.

